# Pandemic telehealth flexibilities for buprenorphine treatment: A synthesis of evidence and policy implications for expanding opioid use disorder care in the U.S

**DOI:** 10.1101/2023.03.16.23287373

**Authors:** Noa Krawczyk, Bianca D. Rivera, Carla King, Bridget C.E. Dooling

## Abstract

Buprenorphine is a highly effective treatment for opioid use disorder and a critical tool for addressing the worsening U.S. overdose crisis. However, multiple barriers to treatment - including stringent federal regulations - have historically made this medication hard to reach for many who need it. In 2020, under the COVID-19 Public Health Emergency, federal regulators substantially changed access to buprenorphine by allowing prescribers to initiate patients on buprenorphine via telehealth without first evaluating them in person. As the Public Health Emergency is set to expire in May of 2023, Congress and federal agencies can leverage extensive evidence from studies conducted during the wake of the pandemic to make evidencebased decisions on the regulation of buprenorphine going forward. To aid policy makers, this review synthesizes and interprets peer-reviewed research on the effect of buprenorphine flexibilities on uptake and implementation of telehealth, and its impact on OUD patient and prescriber experiences, access to treatment and health outcomes. Overall, our review finds that many prescribers and patients took advantage of telehealth, including the audio-only option, with a wide range of benefits and few downsides. As a result, federal regulators—including agencies and Congress—should continue non-restricted use of telehealth for buprenorphine initiation.

## Introduction

COVID-19 struck the United States (U.S.) during an overdose crisis that has claimed over one million lives.^1^ Limited access to highly effective medications for opioid use disorder (MOUD) has long hampered their success as a countermeasure to overdose deaths.^2,3^ Buprenorphine, a partial opioid agonist that can be prescribed by non-specialty providers, is a particularly advantageous MOUD, given its safety profile and its strong protection against overdose risk.^4^ Despite increased availability of buprenorphine over the past two decades,^2^ a vast amount of unmet treatment need persists.^5^

One reason for this gap is that buprenorphine is highly regulated. Until recently, buprenorphine prescribers required a special “X-waiver,” which demanded additional paperwork, training, and prescribing limits.^6^ These barriers left vast regions of the U.S. – especially rural counties – without buprenorphine prescribers.^7^ Moreover, the Ryan Haight Online Pharmacy Consumer Protection Act of 2008 prohibits internet-based prescribing of controlled substances like buprenorphine. The Drug Enforcement Administration (DEA) implemented this statute with regulations that define a “valid prescription” as one made after “at least one in-person medical evaluation.”^8^ Another U.S. regulator, the Substance Abuse and Mental Health Services Administration (SAMHSA) required patients to undergo a physical evaluation before admission to an opioid treatment program (OTP) for buprenorphine treatment.^9^

In response to the COVID-19 public health emergency (PHE), federal regulators issued numerous policy changes that significantly impacted the opioid use disorder (OUD) treatment landscape.^10^ On March 16, 2020, DEA permitted controlled substances, including buprenorphine, to be prescribed via telemedicine (defined as a two-way audio-visual connection) without a prior in-person visit.^11^ On March 31, 2020, DEA relaxed this policy further to allow an audio-only connection.^12^ In April 2020, SAMHSA issued comparable telehealth prescribing guidelines for OTPs.^13^

But in early 2023, the Biden administration signaled the end of the PHE, raising concerns about the pandemic telehealth flexibilities for buprenorphine. SAMHSA recently issued a proposed rule that would incorporate its April 2020 policy into its regulations for OTPs.^14^ DEA also recently proposed to retain some aspects of its pandemic telehealth policy,^15,16^ and Congress has considered proposals to better integrate telehealth into OUD treatment. As Congress and federal agencies consider policy options, they can leverage extensive research to make evidence-based decisions. In this review, our team of public health and regulatory experts synthesizes the evidence on the impact of pandemic buprenorphine telehealth^a^ flexibilities on patient and prescriber experiences and health outcomes. We then interpret the findings for policymakers.

## Methods

### 1. Search strategy

We searched for peer-reviewed studies published between March 1, 2020 and November 15, 2022 focused on measuring the effects of pandemic buprenorphine telehealth flexibilities. We adapted the search strategy from a related review on the impact of pandemic regulatory changes on methadone treatment.^17^ We searched PubMed and PsycInfo with combinations of the following terms: COVID-19, pandemic, buprenorphine, telehealth, opioid treatment program/OTP, opioid-related disorder, and opiate substitution treatment (see Supplemental Table 1 for full search strategy). We also reviewed reference lists from included articles for relevant studies not identified by the database search.

**Table 1.** Characteristics of Included Studies.

| **Study Design** | **n (%)** |
| --- | --- |
| Observational outcomes study | 22 (53.66) |
| Qualitative study | 10 (24.39) |
| Closed-ended survey | 10 (24.39) |
| **Setting of Buprenorphine Treatment** |  |
| Physician Office-Based | 11 (26.83) |
| Virtual Clinic | 4 (9.76) |
| Opioid Treatment Program | 3 (7.32) |
| Syringe Service Program | 4 (9.76) |
| Mobile Clinic | 2 (4.88) |
| Detention Center | 1 (2.44) |
| Multi-Setting | 13 (31.71) |
| Other/Not specified<sup>i</sup> | 5 (12.2) |
| **U.S. Region** |  |
| Northeast | 14 (34.15) |
| South | 6 (14.63) |
| Midwest | 1 (2.44) |
| West | 4 (9.76) |
| Multistate | 16 (39.02) |

<sup>i</sup>Other settings included private and public addiction treatment clinics,<sup>55</sup> outpatient setting,<sup>31</sup> Veterans Health Administration<sup>36</sup> low-barrier, outpatient OUD treatment programs<sup>50</sup>

### 2. Screening and data extraction

We included articles that were: (1) English-language and U.S.-based, (2) original research, and (3) measuring the role or effect of the pandemic buprenorphine telehealth flexibilities. Unlike previous reviews of telehealth for buprenorphine, ^18–21^ we focused exclusively on articles reporting outcomes after the onset of the COVID-19 pandemic and related regulatory changes. Using Covidence, a systematic review tool,^22^ we removed duplicates, screened titles and abstracts, and reviewed the full-text to assess eligibility based on inclusion criteria. The study team conferred to select the final list of articles and then extracted findings based on five research questions deemed to have policy relevance: How the new flexibilities for buprenorphine telehealth provision (1) were implemented in practice; (2) influenced perceptions and experiences of buprenorphine patients and (3) buprenorphine prescribers^b^; (4) affected utilization and retention in treatment, and (5) affected adverse outcomes such as illicit drug use, treatment non-adherence, diversion, and overdose.

### 3. Synthesis of findings

Our team reviewed and synthesized findings related to each of the research questions, considering the different samples, study designs, analytic methods used, and limitations and strengths of each study. We then assessed implications of the findings in terms of potential regulatory or legislative change.

## Results

### 1. Characteristics of reviewed studies

The search resulted in 255 articles, of which 41 met inclusion criteria (Figure 1).

**Figure 1.**
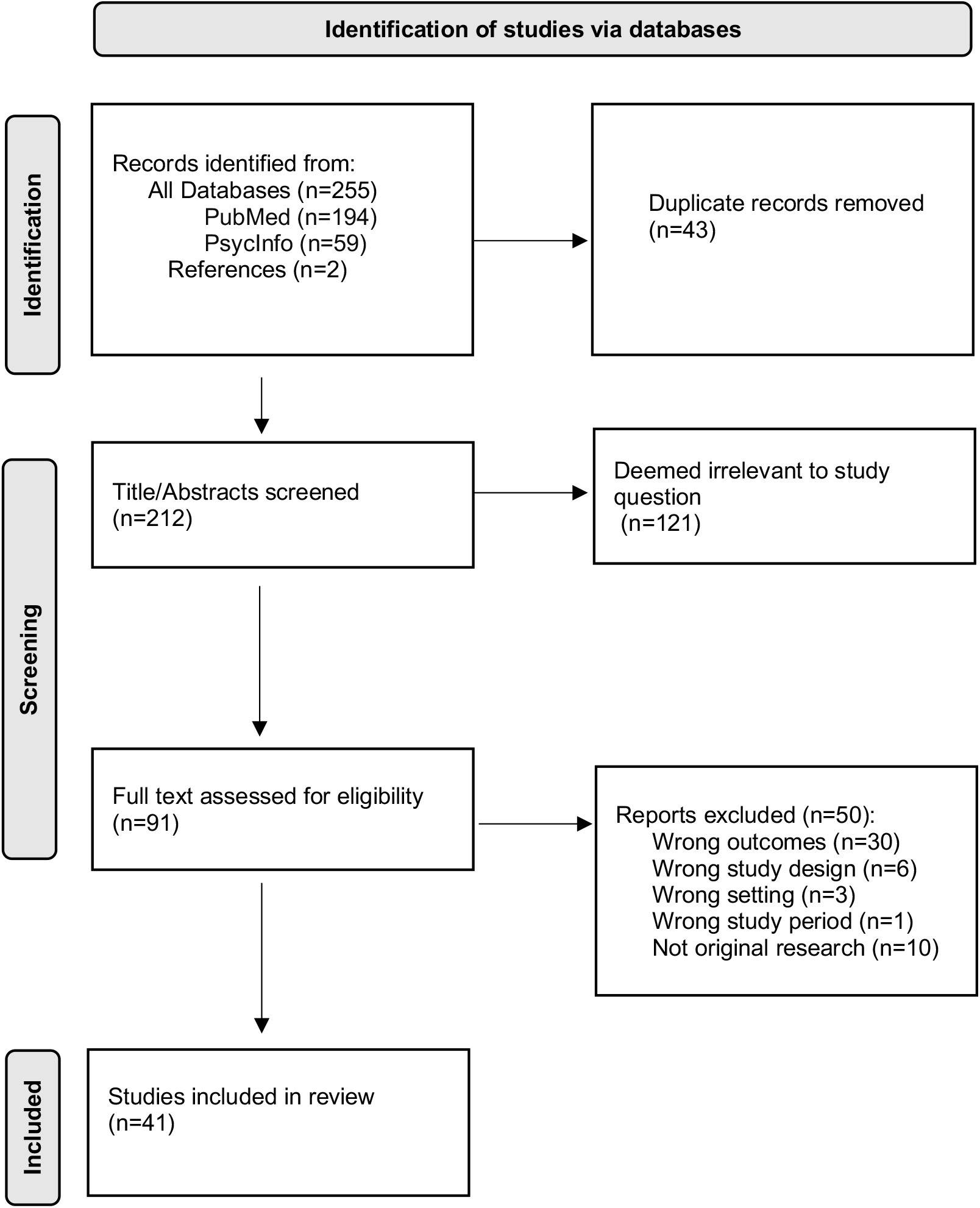
Studies considered and selected in the narrative review.

Characteristics of included articles are summarized in Table 1, with detailed outcomes of each study available in Supplemental Table 2. Most studies were observational outcome studies (N=22 (54%)) and were conducted across multiple treatment settings (N=13, 32%) and multiple U.S. states (N=16, 39%). We include a subset of direct quotes from qualitative research studies conveying patient and prescriber experiences (Table 2). Table 3 summarizes findings on each of the five research questions, described in detail below.

**Table 2.**
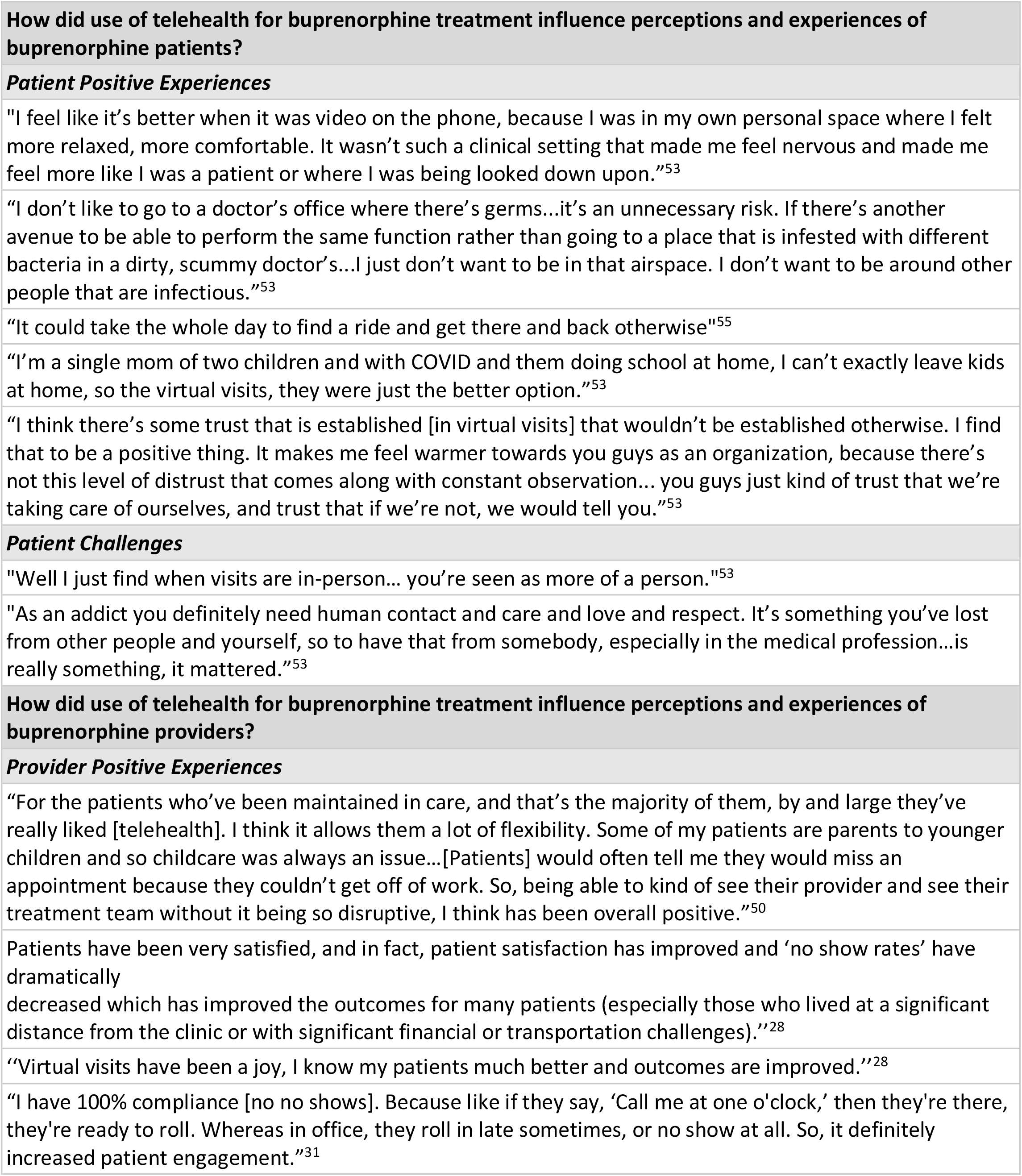
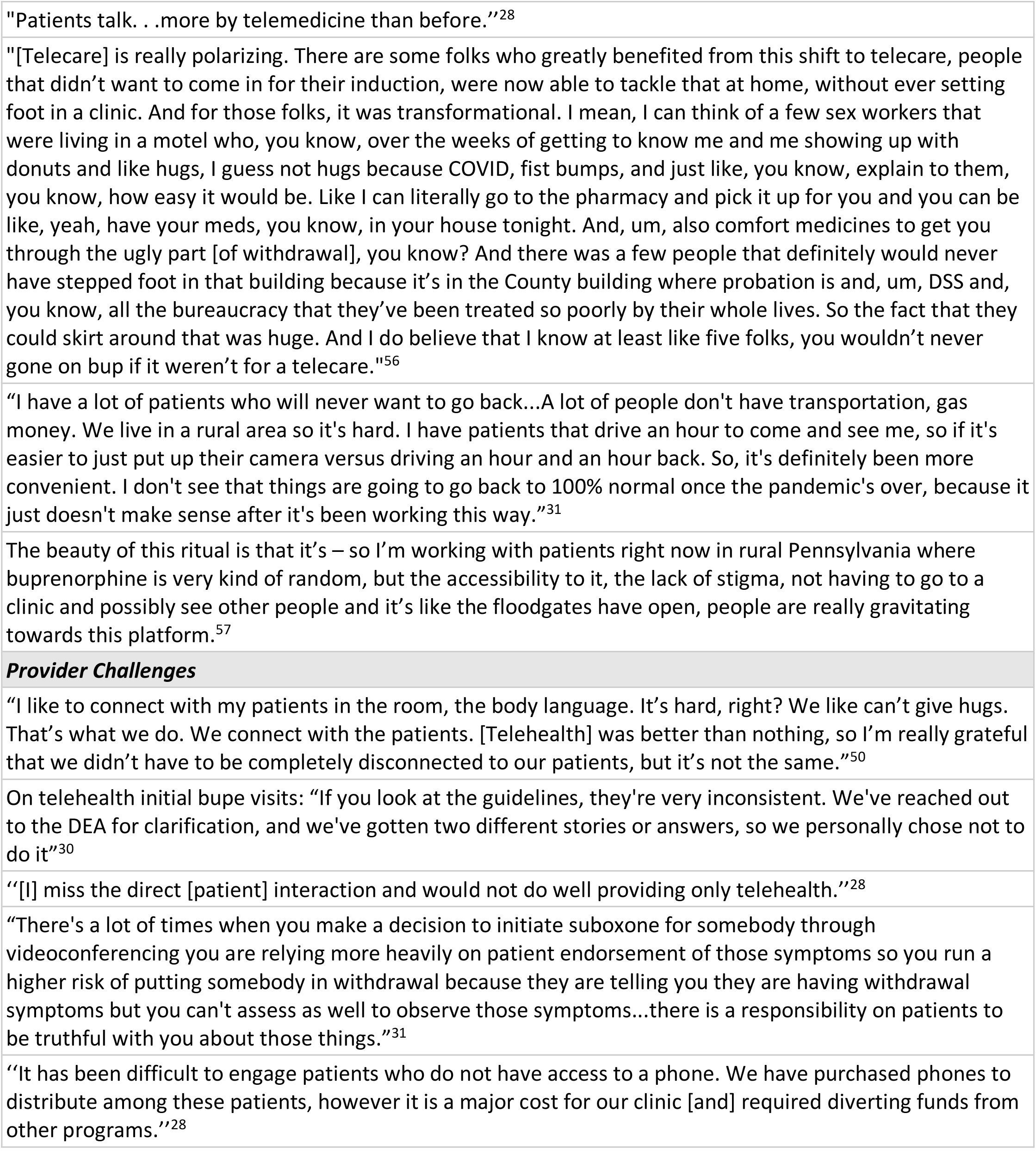
Selected quotes from qualitative studies capturing patient and provider experiences.

**Table 3.** Summary of evidence by policy-relevant question of interest.

|  |
| --- |
| **1. How did use of telehealth for buprenorphine treatment become implemented?** |
| A. Uptake of telehealth for buprenorphine treatment increased following the pandemic regulation changes<sup>23–36,51</sup> |
| B. Initiation of buprenorphine via telehealth was feasible using both video and audio-only technologies, and was successful at engaging patients with health and socioeconomic challenges.<sup>36–42,44–48</sup> |
| C. The shift to telehealth was often accompanied by several practice modifications to support continued patient engagement.<sup>28–30,32,37–39,41,45,49–52</sup> |
| **2. How did use of telehealth for buprenorphine treatment influence perceptions and experiences of buprenorphine patients?** |
| A. Patients reported positive experiences with receiving buprenorphine via telehealth, noting it allotting greater ease and flexibility, reduced burden and stigma, and improved autonomy.<sup>24,28,53–55</sup> |
| B. Some patients expressed challenges with telehealth, including a lowered sense of connection, and ongoing hurdles in accessing medications through remote visits.<sup>53,55,56</sup> |
| **3. How did use of telehealth for buprenorphine treatment influence perceptions and experiences of buprenorphine providers?** |
| A. Many prescribers expressed positive experiences with telehealth, noting benefits to patient care and reduced barriers for initiating and continuing treatment.<sup>28–32,50,51,56–58</sup> Prescribers often preferred video technologies over audio only, but believed audio services could be more accessible to patients.<sup>31,35,51</sup> |
| B. Prescribers noted some challenges and drawbacks of telehealth visits relative to in-person care, including lack of access to technologies, reduced connections and contact with patients, and costs associated with remote visits.<sup>26,28,30–32,50,51,55,58</sup> |
| **4. How did use of telehealth for buprenorphine treatment impact utilization and retention in buprenorphine treatment?** |
| A. Population-based rates of buprenorphine utilization remained relatively stable pre and post-pandemic, but rates of initiation varied across care settings and patient samples.<sup>23,41,59,60</sup> |
| B. Telehealth flexibilities were generally associated with equal or improved retention in care.<sup>36,37,44,46,47,52,61–64</sup> |
| **5. How did use of telehealth for buprenorphine treatment impact adverse outcomes such as illicit drug use, treatment non-adherence or diversion, and overdose?** |
| There was limited assessment of illicit drug use, treatment non-adherence, diversion or overdose risk, but findings did not indicate major concerns around these adverse events.<sup>38,39,48</sup> |

### 2. Synthesis of findings

#### 2.1 Uptake of Telehealth Buprenorphine

Many studies assessed implementation of telehealth regulations by analyzing changes in use of telehealth among patients in buprenorphine treatment. In a study of commercially insured and Medicare Advantage enrollees, telehealth visits for OUD patients filling medications (primarily buprenorphine) increased from 0.52% in week 1 of the pandemic (March 1) to 27.33% in week 13.^23^ A survey conducted in the early months of the pandemic sampling people who use drugs found that among 27 people receiving buprenorphine, 77.8% reported more visits by phone or video.^24^ Some specifically assessed the extent of uptake of telehealth to initiate^c^ buprenorphine, which DEA regulations effectively prohibited before the pandemic. A study using Optum commercial and Medicare Advantage claims found that 14% of 2,703 patients who initiated buprenorphine between April 2020-April 2021 did so via telehealth.^25^

Some assessed the implementation of telehealth regulations by measuring uptake among buprenorphine prescribers and programs. While there was variability across studies, all reported increased use of telehealth for buprenorphine continuation and most for initiation, although often to a lesser magnitude.^26–31^ For example, one survey of 602 buprenorphine prescribers in late 2020 found that 55.8% of all past-month visits and 33.9% of past-month initiation visits were delivered via telehealth.^26^ Another survey found that telehealth utilization rose during the early COVID-19 era (29% pre–COVID-19 to 66% post) and that more prescribers exclusively used telehealth (47%) than in-person visits (33%) for initiation.^29^ Other surveys and interviews of providers similarly found significant uptake of telehealth for buprenorphine among prescribers.^27,28,30,31^

Trends in uptake of telehealth varied across different types of programs offering buprenorphine: A survey of 57 primary care practices found that 92.3% were conducting initiation visits via telehealth,^32^ and a survey of 325 syringe service programs (SSPs) found that a quarter were initiating buprenorphine via telehealth.^33^ In a survey of OTP directors in Pennsylvania, 40% (8) offered video appointments and 50% (10) offered telephone appointments for buprenorphine follow-up visits, but only 25% (5) and 10% (2) respectively did so for initiation. In a survey of local health departments, 72.2% reported their MOUD programs increased telehealth options for OUD.^34^ In a survey distributed by SAMHSA-funded Addiction Technology Transfer Centers, 48% of substance use programs used telephone-based services for buprenorphine, and 57.1% of used video-based services.^35^

A few studies assessed prescriber characteristics associated with uptake of telehealth. One survey found that prescribers more likely to use telehealth for initiation were younger; of emergency medicine specialty vs. primary care; were not in an office-based solo practice; prescribed buprenorphine to more than four patients a month; reported their practice setting closed during the COVID-19 emergency; and had prescribed buprenorphine remotely to established patients prior to COVID-19.^27^ Another found that prescribers were more likely to use telehealth as their prescribing capacities increased, with 50%, 63%, and 71%, using telehealth at patient capacities of 30, 100, and 275, respectively. Addiction medicine physicians used telehealth at higher rates (75%) than physicians from other specialties (61%-63%).^29^ Some studies assessed whether any patient factors were associated with receiving buprenorphine via telehealth. In a study of Medicare Advantage enrollees, younger patients and those in counties with higher median household income were more likely to initiate treatment via telehealth vs. in-person.^25^ On the other hand, in a study of Veterans Health Administration (VHA) patients, younger patients were less likely to have telehealth visits but were more likely to use video versus telephone-only when they did have a telehealth visit.^36^ In a sample of individuals receiving buprenorphine via a mobile health clinic, women and white individuals were more likely to be initiated via telehealth vs. in person.^37^

#### 2.2. Feasibility of Telehealth Buprenorphine

Multiple studies found that telehealth successfully engaged buprenorphine patients with various health and socioeconomic challenges. This included veterans,^36^ individuals experiencing homelessness,^38,39^ individuals with co-occurring psychiatric needs,^38^ individuals with criminal justice involvement,^38^ individuals from rural areas,^41^ individuals who inject drugs,^42^ and individuals from minoritized racial/ethnic groups.^38^ Multiple studies analyzed patient data from ‘low-threshold’ buprenorphine treatment programs, which aim to eliminate as many barriers as possible to accessing care.^43^ Many such programs offered telehealth programs out of mobile vans,^37,44^ SSPs,^40,42,45^ and COVID-19 isolation sites^39^ to provide continued access to buprenorphine initiation and continuation during the pandemic.^38,46^ These programs reported high acceptability and feasibility of patient engagement exclusively via telehealth.^38,39,46,47^

Studies of low-threshold programs noted the importance of offering telephone-only options for patients who did not have access to video technologies.^47^ A few studies thus assessed the feasibility of buprenorphine treatment via telephone, without video technology. An assessment of four telehealth programs across four states found high acceptability of telehealth initiation, with over half of the visits via telephone.^47^ Another study of one buprenorphine practice found that 94.6% of 75 patients had at least one documented telephone-only visit.^48^

#### 2.3. Practice Modifications to Support Telehealth

Many studies explored other changes to MOUD service delivery implemented to support the shift to telehealth-based buprenorphine. Some mentioned the implementation of home-initiation protocols, either via staff education^39^ or use of support technologies such as the “BUP Home Induction”® app.^45^ A program located in a jail described having staff monitor buprenorphine intake for patients who met with a prescriber via telehealth.^49^ Programs also used check-in calls to support patients’ in the transition to telehealth and other difficulties during the pandemic.^37,38^ Others offered patients the option to access remote technologies from a clinic to communicate with a prescriber if they did not have access to personal technologies for telehealth visits.^50^ Others shifted psychosocial services to virtual platforms,^51^ discontinued or required less frequent drug screening,^28,29,32,37,41,51,52^ or required less frequent psychosocial service utilization.^29,51^ Some practices offered longer days-supply to reduce burden,^29,32,37,51^ while others offered shorter days-supply to encourage more frequent checkins.^30^

#### 2.4. Patient Positive Experiences with Telehealth

Many studies found that patients highly supported a telehealth option. They valued that telehealth facilitated initiation and supported access to MOUD throughout the pandemic.^53,54^ Additionally, telehealth reduced transportation and other geographic barriers,^28,55^ improveed patients’ ability to serve as caretakers^53^ and helped them better cope with opioid withdrawal.^53^ Some described virtual visits as less stigmatizing, more comfortable, and fostering self-empowerment and mutual respect between patients and providers.^53^ Some specifically emphasized the importance of having a choice in treatment modality (in-person vs. telehealth).^53^ In a community-based survey that asked about buprenorphine telehealth experiences, 92.8% of participants said that they had the internet/phone connection they needed, 88.6% said they got clear instructions about how to connect, and 84.5% said their care was going pretty well.^24^

### 2.4. Patient Challenges with Telehealth

Despite overall support for telehealth flexibilities, some patients expressed experiencing challenges. They noted that remote visits did not allow them to connect as well as in person, including the loss of tactile connections, which could lead to isolation.^56^ Some noted that inperson care was helpful when experiencing the physical effects of drug use or withdrawal, and that it facilitated meeting their health care needs in one place.^53^ Some patients perceived virtual visits as less serious or easier to delay.^53^ Some patients also reported experiencing unfair treatment by clinic staff via phone^56^ and bureaucratic hurdles at the pharmacy.^55^

### 2.5. Prescriber Positive Experiences with Telehealth

Buprenorphine prescribers generally supported the ability to prescribe buprenorphine via telehealth, with most expressing few to no challenges.^27,28,30,31,57^ Some even noted that they observed better engagement and retention in treatment.^32^ Other observed benefits to patient care included having a better understanding of patients’ lives by being able to see into their homes; reduced burden of patient surveillance from fewer in-person visits and urine drug screens; reduced crowding and exposure to COVID-19; reduced burden of seeing people or places associated with past drug use or trauma; and reduced transportation challenges for those who live in rural areas or have competing responsibilities like work and childcare.^28,31,50,56– 58^ In addition, some described the ability to provide initiation via telehealth as critical to engaging new patients with ambivalence or fluctuating motivation for treatment by allowing for initiation when a patient was ready.^50^

Many prescribers support temporary telehealth flexibilities becoming permanent,^29,30,50^ including one survey that found 85% of 971 prescribers in support.^29^ Some mentioned that reinstating the in-person requirement would be detrimental and prevent patients from accessing needed treatment.^30^ Many specifically mentioned maintaining hybrid models that allow in-person or telehealth care flexibility depending on provider and patient needs and preferences.^30,50^ Some prescribers mentioned the benefit telehealth afforded them personally, such as workplace flexibility^50^ and improved clinic operation efficiency.^31^ Many noted the need to secure equal reimbursement for telehealth services.^30,51^

Some studies assessed factors influencing whether and when prescribers preferred in person vs. video or audio visits. In a study of SUD service providers, respondents preferred video to telephone for most SUD services, but believed audio services were more accessible.^35^ Some prescribers noted prioritizing telehealth for patients at high risk for COVID-19 morbidity or living far away,^31^ while preferring in-person visits for patients who were new, had recently relapsed, had a history of medication non-adherence, lacked devices to support telehealth, were homeless, or had comorbid conditions that required a physical exam or laboratory testing.^31^ Prescribers noted using the telephone for patients with limited computer literacy, or who lacked devices, disliked video, or experienced technical problems with video visits.^31^

### 2.6 Prescriber Challenges with Telehealth

While prescribers generally supported telehealth practices, many noted challenges or drawbacks, and some preferred telehealth not replace in-person care for an extended period.^31^ Noted challenges included inability to rely on clinical tests to monitor treatment adherence,^58^ patient distractions while on video,^58^ inadequate technology among patients or clinics,^51^ and issues with pharmacy dispensing following telehealth visits.^55^ Some felt uncomfortable caring for new patients via video visits,^26^ or had difficulty adapting to the reliance on technology, reduced communication across care teams, and potential for increased liability.^28,32^ Others mentioned hesitancy due to a lack of regulatory clarity,^30^ financial challenges due to lower reimbursement for telephone-only visits, or cost challenges related to purchasing technologies.^30^

Other reasons noted for preferring in-person visits was to gather “tangible and intangible” information^50^ or better engage and connect with patients.^30,50^ Some said they thought patients preferred in-person visits^32^ or benefitted more from them, especially when patients had co-occurring mental health needs.^58^ Some mentioned the difficulty of observing specific physical symptoms of opioid withdrawal via telehealth, detecting “lies” about drug use or diversion,^31^ or believed telehealth could increase diversion and other risks.^51^ Some mentioned difficulty balancing patient preferences with clinician perceptions of patient “stability”^50^ and shifting from “paternalistic” standards of care such as frequent drug screens.^58^ Some of these preferences were associated with provider characteristics. For example, in one study, prescribers working in an OTP were less likely than prescribers working in other settings to prefer telehealth visits.^28^ In a study of buprenorphine prescribers, those with fewer OUD patients were less likely to report feeling comfortable treating patients via telehealth.^26^

### 2.7. Telehealth Impact on Buprenorphine Utilization

Multiple studies compared buprenorphine utilization and initiation rates before and after pandemic flexibilities were implemented. However, they did not always distinguish whether telehealth was involved. One study in a family medicine clinic reported that total MOUD visits increased during the pandemic (436 pre vs. 581 post, p < 0.001), while new patient visits remained constant (33 pre vs. 29 post, p = 0.755).^41^ A study of Texas prescription records found that in the 90 days after the COVID-19 national emergency declaration, 36,225 patients filled buprenorphine prescriptions, up from 30,013 in the preceding 90 days (p < .001). While the number of new buprenorphine patients increased from 4,291 to 13,604 (p < .001), the number of existing buprenorphine patients declined from 25,722 to 22,621 (p < .001).^59^ Another study of Wisconsin Medicaid claims found buprenorphine prescriptions remained stable at about 28% of eligible beneficiaries with OUD leading up to and during early months of the pandemic.^60^ A study using Optum commercial and Medicare claims found no decrease in medication fills or visits among patients receiving MOUD in the first three months of the pandemic. However, they did find that fewer individuals initiated OUD medications.^23^

### 2.8. Telehealth Impact on Buprenorphine Retention

Other studies aimed to assess the impact of the new flexibilities on patient retention. A study of patients in one urban buprenorphine treatment program found a not statistically-significant increase in patient “show rate” from 74.1% (n = 497/671) for prior routine in-person care to 91.7% (n = 166/181) for telehealth visits (p=.06).^61^ In another study of 81 patients at a community clinic, those referred during the pandemic were more likely to be in treatment at 90 days compared to those referred before the pandemic, (68.0% vs. 42.9%, p < 0.05).^52^ Another study assessed 6-month discontinuation among patients receiving buprenorphine from an SSP and found that, compared to patients receiving care prior to the pandemic, those receiving care during the pandemic (either telehealth or in-person) had a lower hazard of discontinuation (respectively, hazard ratios with confidence intervals (CI) of 0.29 (95% CI 0.18, 0.47) and 0.49 (95% CI 0.31, 0.77)), with greatest retention among those receiving care via telehealth.^62^ A study of patients receiving buprenorphine at a rural federally-qualified health center found similar 3-month treatment retention rates pre- and post-COVID-19.^63^

Some feasibility studies of telehealth-based buprenorphine programs established during the pandemic also reported rates of engagement or retention. While these standalone assessments lacked a comparison group, most reported high rates of continued treatment engagement (60% or higher) in the 30 days following the initial prescription.^44,46,47,64^ One study found high levels of treatment retention (>80%) at one year among patients who initiated or continued treatment via telehealth through a mobile van program.^37^ Finally, one study, among a sample of VHA patients who initiated buprenorphine following COVID-19–related regulatory changes, found higher odds of retention among those who had telehealth visits compared to those who had only in-person visits (odds ratio (OR) 1.31; 95% CI, 1.12-1.53).^36^ However, among those who received telehealth, odds of retention was higher among patients who had video visits than only telephone visits (OR: 1.47; 95% CI, 1.26-1.71).^36^

### 2.9. Telehealth Impact on Adverse Outcomes

No studies in our review empirically studied the impact of these flexibilities on overdose, illicit drug use, or buprenorphine diversion. However, two studies assessed whether there were any adverse events associated with telehealth initiation: A study of 12 patients initiated on buprenorphine in a COVID isolation site reported no serious adverse events,^39^ and a study of 199 patients who accessed a virtual buprenorphine program reported few adverse events, including 21 patients who reported withdrawal during induction, four musculoskeletal pain, and one overdose (not necessarily involving buprenorphine).^38^ Only one study assessed buprenorphine adherence by reviewing toxicology records of 21 patients from an office-based treatment program and found a change in positive buprenorphine screens from 92% of patients pre-pandemic to 76% post-pandemic. However, no tests were performed to assess the statistical significance of this change, and no data was reported on what percent of patients were receiving telehealth.^48^

## 3. Discussion and Policy Implications

The wealth of published evidence around the use of telehealth for buprenorphine treatment during the COVID-19 pandemic helps inform important and time-sensitive policy considerations. Following the recent removal of the federal X-waiver requirement for buprenorphine prescribing, questions about the mode of delivery become even more central to expansion efforts for buprenorphine treatment going forward.^65^ Until the pandemic, DEA regulations^66^ made it unlawful for prescribers to use telehealth to initiate patients on buprenorphine. A major finding of our review is that once regulators cleared the way for telehealth initiation, many prescribers and patients took advantage of this new mode of care delivery, including some who might find in-office care to be out of reach for any number of factors. Telehealth visits rapidly increased across various care settings for initiating and continuing buprenorphine treatment. As a result, allowing telehealth likely strongly contributed to relatively stable levels of buprenorphine usage after the pandemic began.

In addition to being feasible, initiation of buprenorphine via telehealth was found to be successful for a variety of patients and generally associated with better retention in care. While the evidence on adverse outcomes was limited, it suggests that telehealth was associated with few-to-no-overdoses, which is consistent with protective effects and known safety profile of buprenorphine.^4^ Since the completion of our review, a recently published study found that despite increasing overdose deaths during the COVID-19 pandemic, there was no increase in the proportion of overdose deaths involving buprenorphine.^67^ Patient experiences were overwhelmingly positive for many reasons including decreased burden, less stigmatized interactions, increased access and freedom, and reduced risks of exposure to COVID-19.

However, such experiences varied across patients, as some did not prefer telehealth to inperson care. This dynamic was generally similar among prescribers, with some enthusiastic about telehealth and hopeful that the flexibilities would continue. Others acknowledged challenges that made telehealth less preferred for them. The preponderance of studies in this review supports the idea that patients and prescribers generally agree that telehealth flexibilities should remain, even while acknowledging that telehealth should not go so far as to replace in-person care for everyone.

The future of telehealth use for buprenorphine is currently in question as the Biden administration announced that the COVID-19 PHE will expire on May 11, 2023. Policymakers should consider the evidence of support for and success of the buprenorphine telehealth experience and act to officially retain telehealth as a mode of initiation for buprenorphine. So far, SAMHSA has proposed a rule to continue to make telehealth available in the OTP setting,^14^ but for all other prescribers, DEA has proposed to extend only a portion of its pandemic flexibility for buprenorphine initiation.^15,16^ The new DEA rule proposes that while prescribers may initiate buprenorphine treatment for 30 days, the patient must make an in-person visit to continue treatment. This is despite prior legal research showing that the DEA possesses ample regulatory authority to keep all of the current regulations, without limits on duration of telehealth.^68^

Findings in our review highlight that a lack of clarity around telehealth regulations may create apprehension to provide these services.^28,30,32^ DEA’s proposed requirement for an inperson visit^15^ may lead to uncertainties around prescribing, limiting uptake. In addition, many studies pointed to the benefits of reaching otherwise hard-to-engage populations via telehealth,^38,39,46^ making it likely that a 30-day mandate for an in-person visit will prevent some at-risk patients from engaging or continuing in care. Given these findings, the DEA should consider removing in-person requirements altogether. Congress could also legislate to make the full pandemic telehealth flexibilities permanent by making audio-only prescribing for buprenorphine clearly acceptable under the Ryan Haight Act. Recent legislation that extended Medicare coverage for telehealth and eliminated the X-waiver demonstrates that there may be a political appetite for such changes that improve access to MOUD.^69^

Even after making these telehealth flexibilities permanent, at least two other policy questions remain. For both, we emphasize that additional access could save lives and suggest a policy approach prioritizing access over other precautionary concerns. Our findings also imply that a treatment landscape without a realistic telehealth option likely means some patients, and perhaps those at greatest risk, will not receive treatment.

First, policymakers should be neutral between video and audio-only telehealth for buprenorphine initiation. DEA’s proposed rule permits audio-only telehealth if the patient does not agree to video, but requires the prescriber to document the patient’s reason for declining video. This implies that the prescriber’s preference is video, when in fact that prescriber’s judgment might be that an audio-only connection is medically acceptable. In reviewed studies, some prescribers preferred video to audio-only modes of communication, but also recognized the access trade-offs of requiring video, a sign of the complexity of this policy choice.

Importantly, our review did not identify substantive evidence that audio-only is less safe or effective than video telehealth. Therefore, as attitudes towards telehealth evolve, it may be wise for policymakers to allow for true flexibility rather than putting a thumb on the scale for video.

Second, the federal government should consider how it might support implementation and adoption of telehealth for buprenorphine initiation. Technical assistance or grant support could help prescribers adapt to new technologies offering secure, HIPAA-compliant, and highquality telehealth services. With this support, prescribers may consider other practices that may be needed to make telehealth a practical option, such as the frequency of urine screening or shifting other support services online.^70^ Our review found that some prescribers, such as older prescribers, those in solo practices, and prescribers treating smaller numbers of OUD patients, lagged behind their counterparts regarding telehealth uptake. Further research is needed to determine if policy interventions, such as training, financial incentives, and more favorable reimbursement rates for telehealth visits,^71,72^ are cost-effective in encouraging telehealth uptake among prescribers. This will be particularly important in expanding these practices to prescribers who were previously not X-waivered but may now consider prescribing buprenorphine. Of course, 100% telehealth uptake is not necessary for a telehealth policy option to improve and save lives.

### Limitations

First, our search was confined to peer-reviewed studies published before November 15, 2022, and may have missed more recent studies. Second, the synthesis and interpretation of the evidence were limited by small sample sizes, variation in settings, time periods assessed, outcomes, and methods used to answer each research question, including solely descriptive approaches. As a result, many of the associations described are not causal. Third, social desirability bias may be present in responses to self-report surveys and interviews. Fourth, the multiple social and structural changes in the pandemic make it challenging to attribute outcomes directly to telehealth regulatory changes. Outcomes should therefore be interpreted in context.

### Conclusion

Under pressure from the pandemic, federal regulators allowed prescribers to initiate patients on buprenorphine via telehealth without first evaluating them in person. Our review finds that many prescribers and patients took advantage of the telehealth option, with a wide range of benefits and few downsides. As a result, federal regulators—including agencies and Congress—should continue the use of telehealth for buprenorphine initiation. Unfortunately, the PHE is set to expire soon and DEA’s proposed rule fails to carry its full pandemic flexibility forward. Given the significant enthusiasm for telehealth and limited-to-no evidence of bad outcomes, DEA should broaden its proposal. If it does not, Congress should act to ensure continuous and low-barrier access to this lifesaving treatment.

## Data Availability

This is a synthesis of research studies so all studies are available in peer-reviewed paper databases

## Acknowledgments and Funding

This work was supported by a grant from the Pew Charitable Trusts. Noa Krawczyk was additionally supported by a grant from the National Institute On Drug Abuse of the National Institutes of Health under Award Number K01DA055758. The content is solely the responsibility of the authors and does not necessarily represent the official views of the National Institutes of Health.

## Conflicts of Interest

Noa Krawczyk provides expert testimony for ongoing opioid litigation

## Footnotes

a We use the term “telehealth” for consistency while some studies may use the term telemedicine for healthcare conducted via audio-only or audio-video communication. We use “telephone” to refer to audio-only (i.e., non-video) telehealth visits.

b We use the term “prescribers” for consistency for what different studies may also refer to as providers or clinicians that prescribe buprenorphine

c We use the term “initiation” for consistency to describe a patient’s initial buprenorphine visit while some studies refer to a is as “induction”

