## Supplement for "Pandemic telehealth flexibilities for buprenorphine treatment: A synthesis of evidence and policy implications for expanding opioid use disorder care in the U.S"

**Supplemental Tables.**

Table 1. Complete search strategy for narrative review

Table 2. Summary table of included studies

**Table 1. Complete Search Strategy for narrative review**

| Database | Strategy | |
|---|---|---|
| | 1 | (“Buprenorphine” AND (“Telemedicine” [Mesh]) |
| | 2 | (“Buprenorphine” OR "Opioid-Related Disorders/drug therapy"[Mesh] OR “Opioid-Related Disorders/therapy"[Mesh] OR "Opiate Substitution Treatment"[MeSH Terms] OR "Opioid-Related Disorders/prevention and control"[Mesh]) AND “Telemedicine” [Mesh] |
| | 3 | (“Buprenorphine” OR "Buprenorphine/therapeutic use"[Mesh] OR "Opioid-Related Disorders/drug therapy"[Mesh] OR “Opioid-Related Disorders/therapy"[Mesh] OR “Opiate Substitution Treatment"[MeSH Terms] OR "Opioid-Related Disorders/prevention and control"[Mesh]) AND “Telemedicine” [Mesh] AND ("COVID-19"[Mesh] OR "Pandemics"[Mesh] OR “SARS-CoV-2” [Mesh]) |
| | 4 | (“Buprenorphine” OR "Buprenorphine/therapeutic use"[Mesh] OR "Opioid-Related Disorders/drug therapy"[Mesh] OR "Opioid-Related Disorders/therapy"[Mesh] OR "Opiate Substitution Treatment"[MeSH Terms] OR "Opioid-Related Disorders/prevention and control"[Mesh]) AND (“Telemedicine” [Mesh] OR “audio-only”) AND ("COVID-19"[Mesh] OR "Pandemics"[Mesh] OR “SARS-CoV-2” [Mesh]) |
| | 5 | (“Buprenorphine” OR "Buprenorphine/therapeutic use"[Mesh] OR "Opioid-Related Disorders/drug therapy"[Mesh] OR "Opioid-Related Disorders/therapy"[Mesh] OR "Opiate Substitution Treatment"[MeSH Terms] OR "Opioid-Related Disorders/prevention and control"[Mesh]) AND (“Telemedicine” [Mesh] OR “audio-only” OR “audio” or “telemedicine” or “telehealth” or “virtual” or “video”) AND ("COVID-19"[Mesh] OR "Pandemics"[Mesh] OR “SARS-CoV-2”[Mesh]) |
| | 6 | (“Buprenorphine” OR "Buprenorphine/therapeutic use"[Mesh] OR "Opioid-Related Disorders/drug therapy"[Mesh] OR "Opioid-Related Disorders/therapy"[Mesh] OR "Opiate Substitution Treatment"[MeSH Terms] OR "Opioid-Related Disorders/prevention and control"[Mesh]) AND (“Telemedicine” [Mesh] OR “audio-only” OR “audio” or “telemedicine” or “telehealth” or “virtual” or “video”) AND ("COVID-19"[Mesh] OR "Pandemics"[Mesh] OR “SARS-CoV-2”[Mesh] or “COVID” or ”pandemic”) |
| PubMed | 7 | ((("COVID-19"[Mesh] OR "Pandemics"[Mesh] OR "SARS-CoV-2"[Mesh] or COVID or pandemic)) AND (("Telemedicine" [Mesh] OR audio-only OR audio or telemedicine or telehealth or telephone or virtual or video OR online))) AND ((opioid related disorders AND (treatment or therapy)) OR Opiate Substitution Treatment OR Buprenorphine) |
| | 1 | covid-19.mp. or exp COVID-19/ |
| | 2 | exp Pandemics/ or exp Severe Acute Respiratory Syndrome/ or sars-cov-2.mp. |
| | 3 | 1 or 2 |
| | 4 | telemedicine.mp. or exp Telemedicine/ |
| | 5 | telehealth.mp. |
| | 6 | exp Online Therapy/ |
| | 7 | (audio-only or audio or telemedicine or telehealth or telephone or virtual or video or online).mp. |
| | 8 | 4 or 5 or 6 or 7 |
| | 9 | 3 and 8 |
| | 10 | buprenorphine.mp. or exp Buprenorphine/ |
| | 11 | exp Drug Therapy/ or exp Treatment/ |
| | 12 | exp "Opioid Use Disorder"/ or "Opioid use disorder".mp. |
| | 13 | 11 and 12 |
| | 14 | 10 or 13 |
| | 15 | 9 and 14 |
| | 16 | 9 and 10 |
| | 17 | 9 and 13 |
| PsycInfo | 18 | 9 and 12 |

**Table 2. Summary table of included studies**

| Author, Year | Study Design | Setting of Care, Location | Study Period (Pre, During, 'Post' Guideline Change) | Sample population | Sample size |
|---|---|---|---|---|---|
| Aronowitz, 2021 | Qualitative study | Other, Philadelphia, Pennsylvania | July - August 2020 | MOUD providers | 22 |
| Barsky, 2022 | Observational outcomes study | Multi-setting, Multistate | Pre: January 1 - March 30, 2020<br>During: April 1, 2020 - April 30, 2021 | Adults with OUD | 3,638 |
| Beetham, 2022 | Closed end survey of practices/perceptions | Office-based, Multistate | July 2020 | MOUD providers | 1,054 |
| Belcher, 2021 | Observational outcomes study | Detention Center, Talbot County, MD | August 15, 2020 - February 15, 2021 | Adults with OUD in jail, awaiting trial or bail | 7 |
| Cance, 2020 | Observational outcomes study | Multi-setting, Texas | Pre: December 15, 2019, - March 13, 2020<br>During: March 14 - June 11, 2020 | Adults with OUD | not specified |
| Castillo, 2020 | Observational outcomes study | SSP, Miami, FL | March 30 - June 8, 2020 | People who inject drugs | 31 |
| Caton, 2021 | Closed end survey of practices/perceptions | Office-based, California | April 20 - May 8, 2020 | MOUD providers | 118 |
| Caulfield, 2021 | Qualitative study | Setting of care not specified, New York | March 2020 - July 2021 | Adults with OUD | not specified |
| Cunningham, 2022 | Observational outcomes study | Office-based, Bronx, NY | Pre: March - August 2019<br>During: March - August 2020 | Adults with OUD | 107 |

| Author, Year | Study Design | Setting of Care, Location | Study Period (Pre, During, 'Post' Guideline Change) | Sample population | Sample size |
|---|---|---|---|---|---|
| Frost, 2022 | Observational outcomes study | Other, Multistate | March 23, 2020 - March 22, 2021 | MOUD providers | 17,182 |
| Harris, 2022 | Observational outcomes study | Mobile Clinic, Baltimore City, MD | March 16, 2020 - March 15, 2021 | Adults with OUD | 150 |
| Hughes, 2021 | Observational outcomes study | Office-based, North Carolina | Pre: January 16 - March 15, 2020<br>COVID transition: March 16 – April 15, 2020<br>During: April 16 - June 15, 2020 | Adults with OUD | 242 |
| Huskamp, 2020 | Observational outcomes study | Multi-setting, Multistate | January - May 2020 | Adults with OUD | 5,988,307 |
| Huskamp, 2022 | Closed end survey of practices/perceptions | Multi-setting, Multistate | November - December 2020 | MOUD providers | 602 |
| Jones, 2021 | Closed end survey of practices/perceptions | Multi-setting, Multistate | June 23 - August 19, 2020 | MOUD providers | 10,326 |
| Kaur, 2022 | Observational outcomes study | Office-based, Pennsylvania | July 1, 2019 - June 30, 2020 | Adults with OUD | Varies by time period analyzed: 309, 327, 360,365 |
| Krawczyk, 2021 | Qualitative study | Multi-setting (Reddit: r/opiates, r/OpiatesRecovery), Multistate | March 5 - May 13, 2020 | Adults with OUD | 300 posts |
| Krawczyk, 2022 | Closed end survey of practices/perceptions | Multiple OTPs, Pennsylvania | September - November 2020 | Clinical directors of OTPs | 47 |

| Author, Year | Study Design | Setting of Care, Location | Study Period (Pre, During, 'Post' Guideline Change) | Sample population | Sample size |
|---|---|---|---|---|---|
| Lambdin, 2022a | Observational outcomes study | SSP, California | May 28, 2020 - March 26, 2021 | Adults with OUD | 114 |
| Lambdin, 2022b | Closed end survey of practices/perceptions | SSP, Multistate | February - June 2021 | Syringe service programs | 295 |
| Lockard, 2022 | Qualitative study | Office-based, Portland, OR | March - April 2021 | Adults with OUD | 19 |
| Lynch, 2022 | Observational outcomes study | Virtual Clinic, Pennsylvania | April 27, 2020 - July 31, 2021 | Adults with OUD | 200 |
| Mattocks, 2022 | Qualitative study | Office-based, Multistate | October 2020 - May 2021 | MOUD providers | 23 |
| Mehtani, 2021 | Observational outcomes study | Virtual clinic, San Francisco, CA | April 10 - May 25, 2020 | Adults with OUD | 19 |
| Molfenter, 2021 | Closed end survey of practices/perceptions | Multi-setting, Multistate | May 15 - August 31, 2020 | Organizations providing SUD services | 457 |
| Nordeck, 2021 | Observational outcomes study | Mobile Clinic, Baltimore City, MD | March 1 - May 31, 2020 | Adults with OUD | 143 |
| O'Gurek, 2021 | Observational outcomes study | Office-based, Philadelphia, Pennsylvania | Pre: January 1 - March 13, 2020<br>During: March 16 - April 30, 2020 | Adults with OUD | 852 visits |
| Rahman, 2021 | Observational outcomes study | Office-based, Massachusetts | March 1st - June 15th, 2020 | Adults with OUD | 75 |
| Saloner, 2022 | Closed end survey of practices/perceptions | Multi-setting, Multistate | August 19, 2020 - January 29, 2021 | Individuals receiving substance use treatment | 316 |
| Samuels, 2022 | Observational outcomes study | Multi-setting, Multistate | March - June 2020 | Adults with OUD | 159 |

| Author, Year | Study Design | Setting of Care, Location | Study Period (Pre, During, 'Post' Guideline Change) | Sample population | Sample size |
|---|---|---|---|---|---|
| Sivakumar, 2022 | Observational outcomes study | SSP, New Haven, Connecticut | March 15 - June 15, 2020 | Adults with OUD | 31 |
| Sung, 2022 | Closed end survey of practices/perceptions; Qualitative study | Multi-setting, Multistate | July - August 2020 | MOUD providers | 797 |
| Swann, 2022 | Closed end survey of practices/perceptions | Multi-setting, Multistate | November 11 - December 23, 2020 | Local health departments | 214 |
| Textor, 2022 | Qualitative study | Other & multi-setting, Multistate | May 2020 - May 2021 | Adults with OUD; MOUD providers; pharmacists | 24 providers and clinical staff; 10 pharmacists; 19 patients |
| Tilhou, 2022 | Observational outcomes study | OTP & Office-based, Wisconsin | Pre-PHE: December 1, 2018 - March 15, 2020<br>Early-PHE: March 16 - May 15, 2020<br>Later-PHE: May 16 - September 30, 2020<br>[PHE=public health emergency] | Adults with OUD | 6,453 |
| Tofighi, 2022 | Observational outcomes study | Virtual Clinic, New York City | March 2020 - March 2021 | Adults with OUD | 99 |
| Treitler, 2022 | Qualitative study | OTP & Office-based, New Jersey | September - November 2020 | MOUD providers | 20 |
| Uscher-Pines, 2020 | Qualitative study | Other, Multistate | April 15 - April 24, 2020 | MOUD providers | 18 |
| Walters, 2022 | Qualitative study | Multi-setting, Northeast | June - October 2020 | Adults ≥ 18 who use/used drugs currently, previously | 13 were currently on buprenorphine, 3 were previously on |

| Author, Year | Study Design | Setting of Care, Location | Study Period (Pre, During, 'Post' Guideline Change) | Sample population | Sample size |
|---|---|---|---|---|---|
| | | | | or never engaged in buprenorphine treatment; MOUD providers, clinic staff, or work at a regulatory agency | buprenorphine, 6 had never been on MOUD; 8 buprenorphine providers |
| Ward, 2022 | Observational outcomes study | Office-based, Philadelphia, Pennsylvania | Pre: September 1, 2018 - March 12, 2020<br>During: March 13 - December 2020 | Adults with OUD | 506 |
| Wunsch, 2022 | Observational outcomes study | Virtual Clinic, Rhode Island | April 2020 - February 2021 | Adults with OUD | 134 |
